# Whole-blood transcriptomic traces of organ pathology and their causal triage

**DOI:** 10.64898/2026.07.31.26359425

**Authors:** Ranjit Kumar Sinha, Kyle Alvarez, Sanju Sinha

## Abstract

Whole blood offers a non-invasive window into organ health, yet it remains unclear which organ pathologies leave a detectable trace in the blood transcriptome, and whether such traces are causal or reactive. Progress has been limited because paired whole-blood profiles and pathologist-graded organ pathology are rarely available together. Here we assembled a pathology-linked benchmark of 59 pathologies across 20 organs in 803 GTEx v10 donors with matched whole-blood RNA-seq and postmortem histology. Because a postmortem cohort’s blood is strongly shaped by age, sex, and the circumstances of death, our framework, *TRACE*, counts a signal only when blood expression predicts a pathology beyond these donor factors. Four pathologies passed, with liver cirrhosis by far the strongest (AUC 0.79). The cirrhosis signature replicated in an independent cohort of living patients (AUC 0.80) and remained specific against severe systemic illness. To separate candidate drivers from reactive markers, we used human genetics: Mendelian randomization linking plasma proteins to liver disease, which recovered established fibrosis drivers, including PAI-1 (SERPINE1), tenascin-C, thrombospondin-2 and nominated further candidates. Together, TRACE provides a resource and a confounder-aware framework for learning which organ pathologies the blood transcriptome can, and cannot, detect, and which of those signals are likely causal.

## Introduction

Most complex diseases involve organs that are difficult to sample in living people, because biopsy is invasive and often impractical. Whole blood is the exception: it is collected routinely, at scale, and non-invasively, which has made it the default substrate for molecular biomarkers. This raises a question that has not been asked systematically: for how many organ pathologies does the whole-blood transcriptome actually carry a detectable trace, and where it does, do those traces reflect processes that drive the pathology or merely react to it? Individual blood-based signatures have transformed diagnosis in specific settings [1, 2], but they have been built one disease at a time, without a common benchmark of which organ pathologies are readable from blood at all.

Three largely separate lines of work have approached organ health from blood, and each stops just short of this question. Plasma-proteomic organ-aging clocks assign circulating proteins to organs and estimate organ-specific biological age at biobank scale [3, 4]; they have reshaped how the field reads organ health from blood, but they are trained on chronological age and validated against diagnosis codes, incident disease, or mortality, never against the pathology of the organ itself in the same person. Blood-to-tissue inference methods predict tissue-specific expression from genotype [5, 6] or from the whole-blood transcriptome [7–9], but treat tissue expression as the endpoint and stop before structural pathology. Proteome-wide Mendelian randomization screens circulating proteins against disease phenomes [10, 11], but anchors on registry diagnoses rather than tissue histology. Two things are therefore consistently missing: a within-person link between a blood signal and pathologist-graded organ pathology, and a causal test applied to the predictive features.

The Genotype-Tissue Expression (GTEx) project is uniquely placed to close this gap, pairing whole-blood RNA-seq with postmortem organ histopathology and germline genotype across hundreds of donors and dozens of tissues [12, 13]. However, there are a few challenges to use GTEx to close this gap. First, the pathology labels are hidden within unstructured notes of pathologist. Even if we solve this problem using recent advancements in natural language processing, using this data for this purpose demands unusual care. This is because death itself reshapes the blood transcriptome: post-mortem interval, agonal state, age, sex, and race dominate donor-level expression variance [14]. A classifier trained on blood without accounting for these factors risks learning the circumstances of a donor’s death rather than the biology of their organ. The meaningful test is therefore not whether blood expression predicts a pathology, but whether it predicts beyond what donor demographics and procedural variables already explain, and, separately, whether the predictive features drive the pathology or follow from it.

Here we introduce TRACE (**T**ranscriptomic **R**eading of **A**ge-associated organ pathologies and **C**ausal **E**valuation), a confounder-aware framework that asks these two questions in sequence across a pathology-linked GTEx benchmark of 59 pathologies spanning 20 organs in 803 donors. We first ask which organ pathologies the whole-blood transcriptome predicts beyond a donor clinical-covariate baseline, and find that most do not: only four pathologies clear the bar, with liver cirrhosis the clearest by a wide margin. We then ask which of the predictive features are causal, using Mendelian randomization to link plasma proteins to liver disease through human genetics; this recovers established drivers of fibrosis, confirming that the signature points to real biology, and nominates further candidates. The contribution is deliberately scoped. This is not a new diagnostic (the cirrhosis signature does not outperform existing clinical liver tests) but a resource and a method for separating organ pathologies that leave a genuine, confounder-independent trace in blood from those that do not, and for triaging that trace into candidate causal and reactive components. Liver serves as the worked example throughout.

## Results

### TRACE: a framework that detects organ pathology in blood, then triages the signal for cause

TRACE rests on a two-part logic: *first detect, then test for cause*. A blood signal counts as organ-pathology signal only if it survives the demographic and procedural variation that pervades a postmortem cohort like GTEx, so a confounder-independent correlation is the entry criterion, not the endpoint. Because such a correlation cannot reveal whether a blood feature drives the pathology or merely reacts to it, TRACE then passes the surviving signal to a genetic causal test that prioritizes the features more likely to be causal. The framework runs in four steps (**Fig. 1**). First, it converts histology slide annotations into donor-level pathology labels, combining GTEx’s structured pathology categories with a natural-language rescue of free-text pathologist notes (**Fig. 1A**, Methods 2). Second, it represents the whole-blood transcriptome by its principal components and removes any component that tracks a donor clinical covariate before that component can enter a model (**Fig. 1B**, Methods 3-4). Third, it trains a classifier for each pathology and asks whether the classifier predicts beyond a covariate-only baseline; pairs that clear a pre-specified bar advance to gene- and pathway-level interpretation (**Fig. 1C**, Methods 3-5). Fourth, it triages the predictive features by Mendelian randomization, separating candidate causal drivers from reactive markers (**Fig. 1D**, Methods 6). The first three steps establish where a trace exists; the fourth asks whether it is cause or consequence.

**Figure 1.**
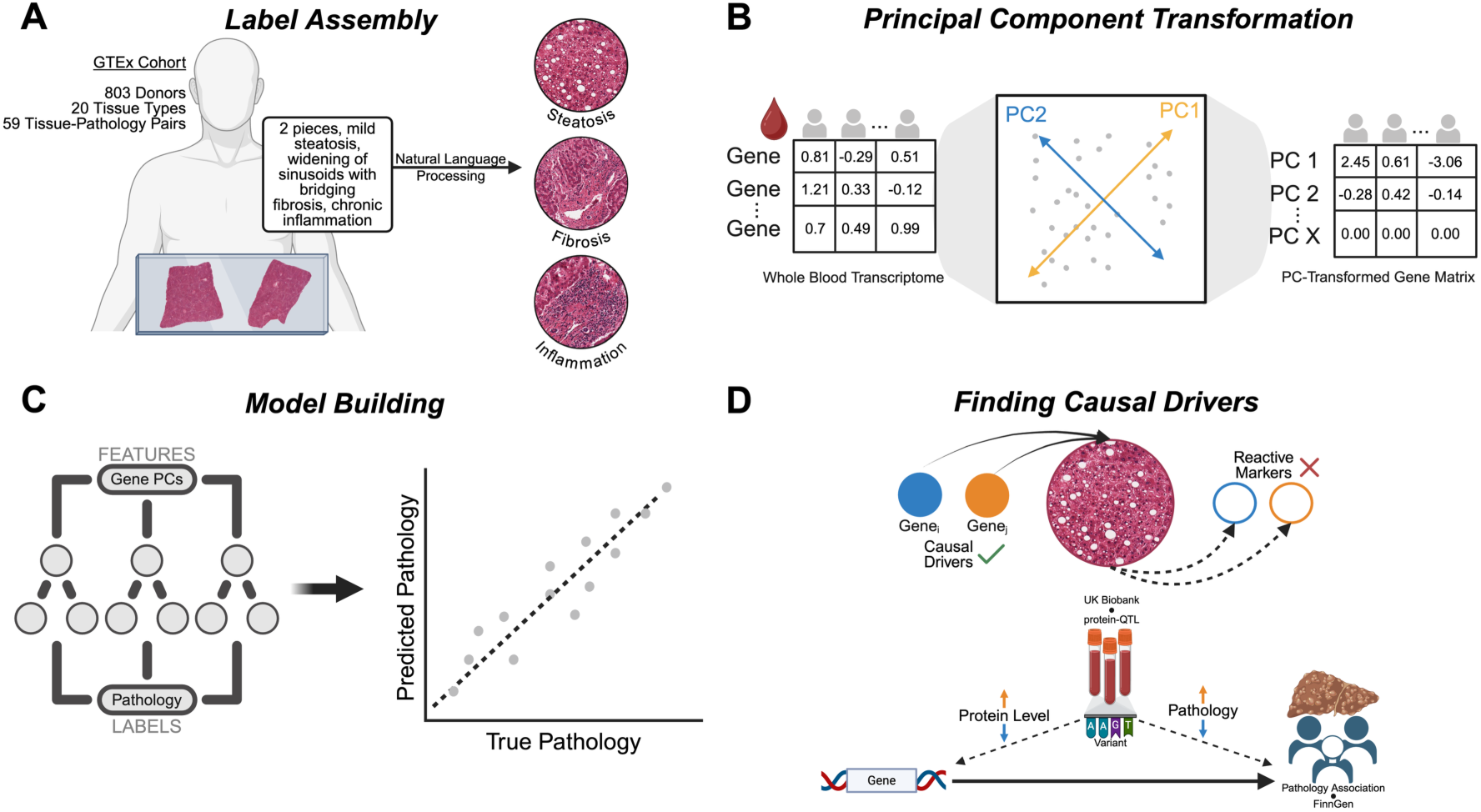
Overview of the TRACE framework. (A) Pathology label assembly, combining structured GTEx histology annotations with natural-language rescue of unstructured pathologist notes. (B) Principal-component transformation of the whole-blood transcriptome with per-fold covariate orthogonalization. (C) Supervised classification against a clinical-covariates baseline, with gene- and pathway-level interpretation of the qualifying pairs. (D) Two-sample Mendelian randomization on pathology-associated blood transcripts, separating candidate causal drivers from reactive markers.

### In GTEx blood, donor demographics and manner of death, not pathology, dominate expression variance

Per-pathology donor counts ranged from 108 to 633 across the 59-pair pool spanning 20 tissue types (Fig. 2A). The cohort’s age, Hardy-scale, and total-ischemic-time distributions are typical of a postmortem sample (**Fig. S1C–E**). To characterize the cohort’s expression structure, we ran principal-component analysis on the 20,000 highest-variance genes. The first two components captured 29.8% and 14.4% of total variance (Fig. S1B), and both encoded procedural rather than pathological signal: PC2 was strongly graded by ischemic time (rho = -0.74) and Hardy scale (rho = -0.67), moderately by age (rho = -0.35), and weakly by sex, while PC1 correlated only weakly with all five covariates (**Fig. 2B**). Because these dominant axes track how a donor died rather than what their organs looked like, every blood model must be defended against them. TRACE does this in two ways: each model is benchmarked against a covariates-only baseline, and, before any component enters a classifier, a per-fold gate drops any principal component whose univariate AUC against a covariate exceeds 0.70 (Methods 4). This gate removed a median of 20 components per pair-fold (range 3-89), concentrated in the low-order components where ischemic-time and Hardy-scale signal is strongest (**Fig. 2C**, **Fig. S2B**).

**Figure 2.**
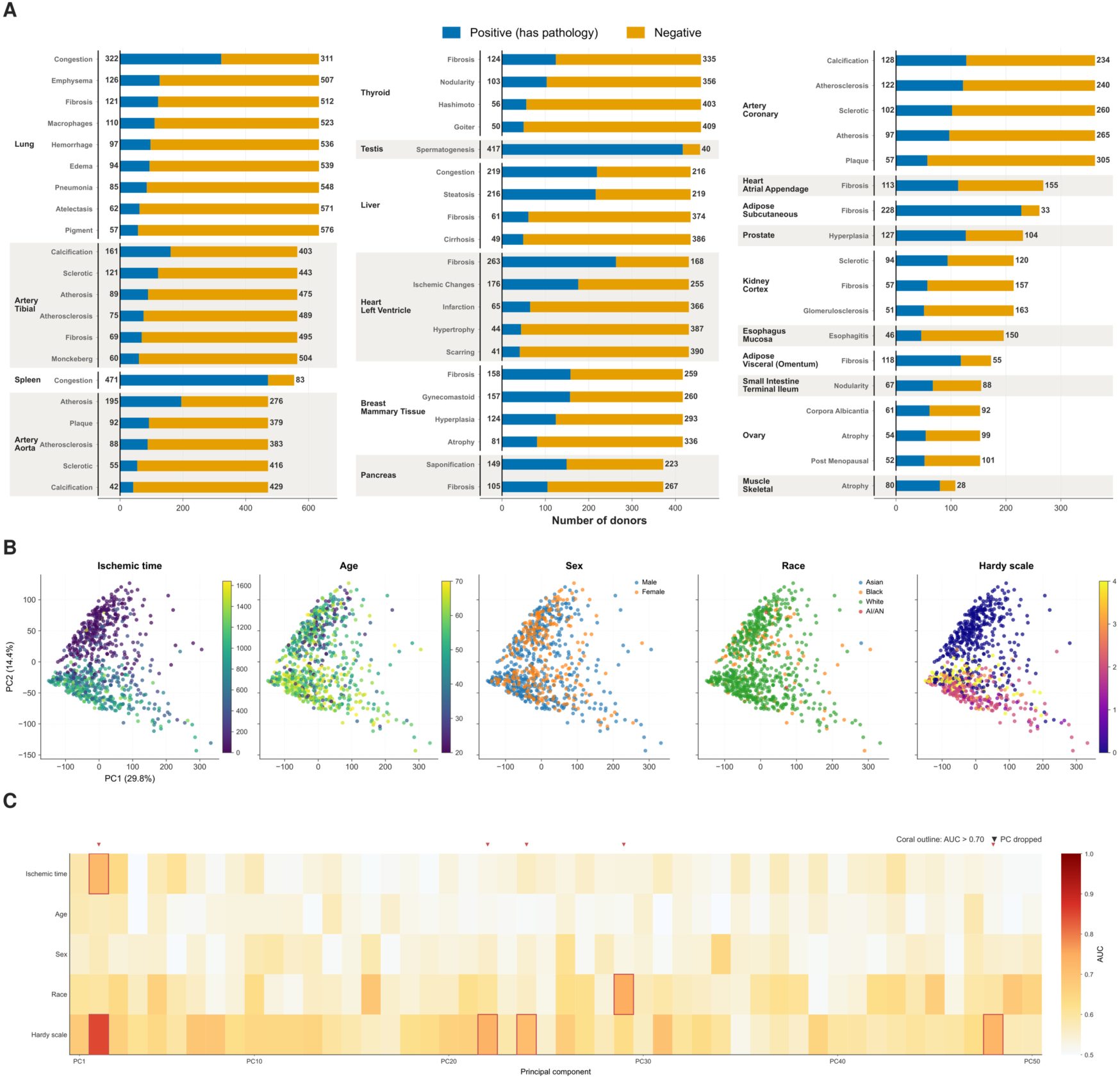
Donor cohort, expression structure, and clinical-covariate control across the 59 GTEx tissue-pathology pairs. (A) Per-pathology donor counts across 59 tissue-pathology pairs spanning 20 GTEx tissue types; pathology-positive donors in blue, pathology-negative in gold; tissues grouped by dashed brackets. (B) PC1 versus PC2 of the whole-blood transcriptome (20,000 highest-variance genes), coloured in turn by five clinical covariates: ischemic time, age, sex, race, and Hardy scale. (C) Per-PC univariate AUC against each of the five covariates, shown for PC1- PC50; coral outlines mark PCs exceeding the AUC = 0.70 threshold, which are dropped by the per-fold covariate-orthogonalization step before entering any pathology classifier.

### Only four of 59 organ pathologies can be traced in blood beyond donor confounders

With the confounder controls in place, we asked the first question: which organ pathologies does the whole-blood transcriptome predict beyond donor demographics, and can learn within in the GTEx cohort? The answer is a minority. Across all 59 pairs, the principal-component random forest exceeded the covariates-only baseline in only four, spanning three tissues (**Fig. 3B-C**; **Fig. S2C-D**, **S3A**): liver-cirrhosis (AUC = 0.79, gain = +0.215), liver-steatosis (AUC = 0.67, gain = +0.148), lung-congestion (AUC = 0.67, gain = +0.053), and small intestine-nodularity (AUC = 0.62, gain = +0.080). These four calls were also the most stable across cross-validation folds: per-fold AUC standard deviation across the full 59-pair pool ranged from 0.02 to 0.16, with all four qualifying pairs below the median fold variance (**Fig. S2E**). This selectivity is itself a result. Because 55 of 59 pathologies produced no confounder-independent signal under an identical procedure, the four that pass are unlikely to be artefacts of the covariates that dominate the cohort. The bar rewards incremental signal over confounders, not absolute accuracy: lung-pneumonia missed at a gain of +0.011 despite a competitive absolute AUC (**Fig. 3C**).

**Figure 3.**
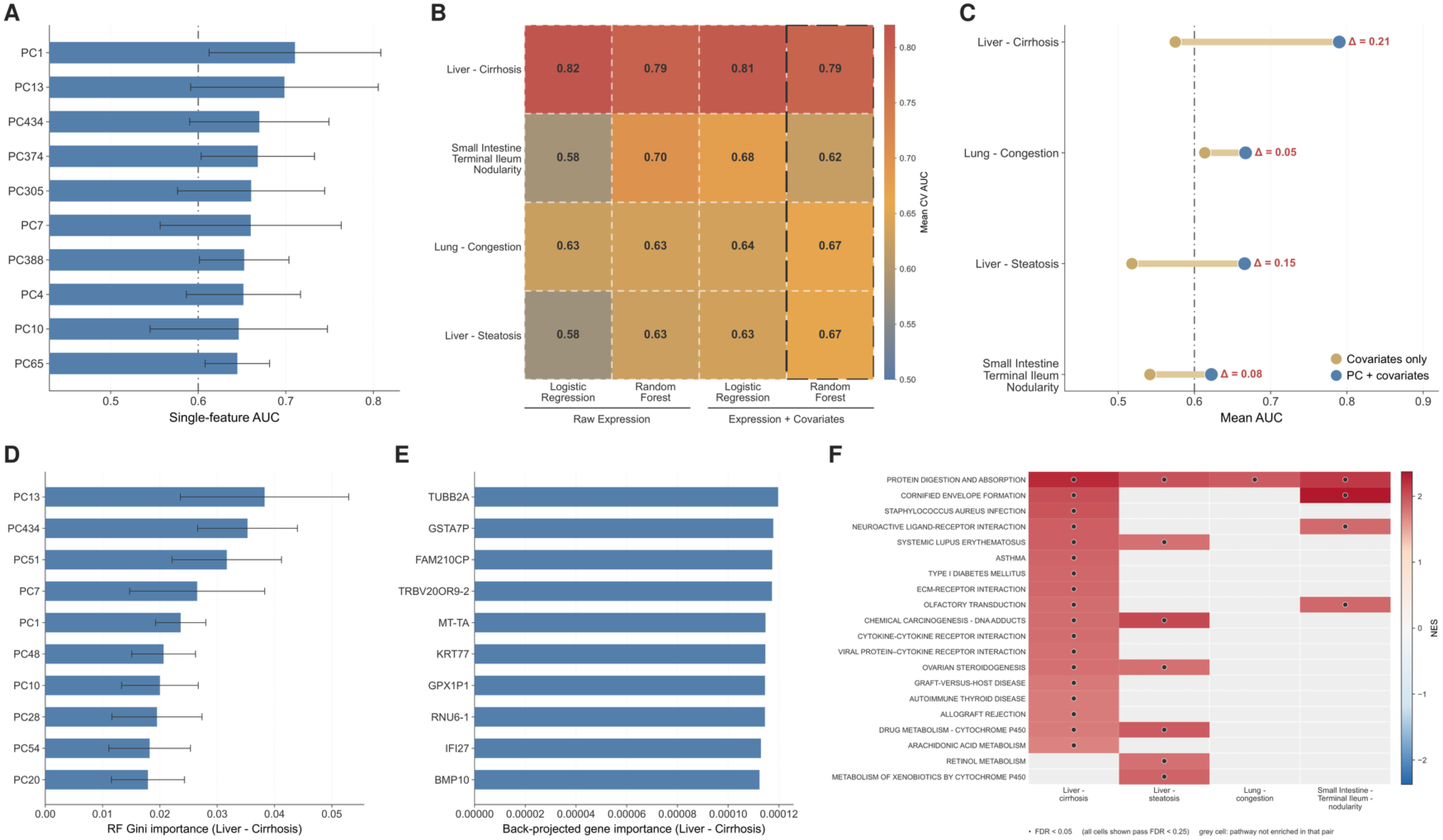
Blood-expression modeling identifies four qualifying tissue-pathology pairs and their gene- and pathway-level interpretation. (A) Top principal components ranked by mean single-feature AUC for liver-cirrhosis; bars show standard deviation across folds. (B) Cross-validated mean AUC across the four qualifying pairs for four model classes (logistic regression and random forest, on raw expression and on PC-transformed expression); the highlighted column marks the best-performing class (PC + covariates random forest). (C) Baseline AUC (clinical covariates only) versus PC-plus-covariates AUC per qualifying pair, with AUC gain annotated. (D) Principal components ranked by random-forest Gini importance for liver-cirrhosis. (E) Top-10 genes for liver-cirrhosis, ranked by PC-derived importance. (F) KEGG 2026 top-20 union pathways across the four qualifying pairs, ordered by liver-cirrhosis NES; cells shaded by normalized enrichment score, black dots mark FDR < 0.05, and all shown cells pass FDR < 0.25.

Liver-cirrhosis stood apart. Its covariates-only baseline was AUC = 0.58, so donor demographics leave most of the cirrhosis signal unexplained, and the full model reached 0.79. The low-dimensional representation lost no information relative to the raw transcriptome: the principal-component random forest matched a gene-level random forest exactly (both AUC = 0.79), with logistic-regression variants comparable (**Fig. 3B**; **Fig. S3B-D**). A single-feature scan showed that a small set of components (PC1, PC13, PC434, PC374, and PC305) each carried most of the signal on its own (**Fig. 3A**). We interpret these components next.

### The liver-cirrhosis blood signature converges on complement, mitochondrial, and matrix-remodeling programs

To read the cirrhosis model at the gene level, random-forest importances over the selected components were back-projected through PCA loadings into a fold-averaged gene ranking (**Fig. 3E**, Methods 4), The principal components the model weighted most (PC13, PC434, PC51, PC7, PC1; **Fig. 3D**) overlapped those that individually carried the signal, indicating the model concentrated its decision on the cirrhosis-informative axes. Agreement between this back-projected ranking and an independent gene-level random forest was rho = 0.56, the highest of any qualifying pair (**Fig. S4B-C**), and the PC ranking was stable across folds (**Fig. S4A**). We further tested the signature-specificity of each pair applied to every other pair (**Fig. S5A**) and utilized the plasma-organ atlas of Oh et al to test cross-organ plasma enrichment (**Fig. S5B)**. We note that a fraction of the highest-importance transcripts are pseudogenes and non-coding loci; because these carry no pathway or protein annotation, all downstream enrichment and Mendelian-randomization analyses were restricted to the annotated protein-coding gene universe (Methods 4).

On that annotated ranking, pre-ranked gene-set enrichment [15] recovered coherent and disease-relevant programs. KEGG 2026 [16] was led by protein digestion and absorption, cornified-envelope formation, and complement-linked immune terms(**Fig. 3F**), and Reactome 2024 [17] by mitochondrial tRNA processing and complement activation (creation of C4 and C2 activators; classical antibody-mediated complement activation) (**Fig. S5C**). The three programs dominate the cirrhosis signature: complement and humoral host-defense [18], mitochondrial RNA processing [19], and epithelial and extracellular-matrix remodeling [20], each with a clear correspondence to established cirrhosis biology.

### Causal triage recovers established fibrosis drivers and replicates in living patients

Circulating biomarker discovery is more likely to be successful when features are causal to pathology. However, above gene rankings are correlational: a protein can rank highly because it drives cirrhosis or simply because it rises in response to the damage. To separate the two, we used a mathematical test called Mendelian Randomization (MR). MR uses large population genetics data to prioritize genes causally related to the pathology. The principle is following: MR uses human genetics as a natural experiment. People inherit small DNA differences that raise or lower a given protein’s level from birth, independent of lifestyle or disease; if those born with genetically higher levels also carry higher liver-disease risk, the protein is more likely a cause than a consequence. Formalized as two-sample Mendelian randomization [21, 22], we linked genetic predictors of plasma-protein levels in UK Biobank [23] to genetic associations with liver disease in FinnGen [24, 25] (Methods 6). Of the 100 most up-regulated cirrhosis transcripts, 15 showed genetic evidence of causing at least one of six liver phenotypes (P < 0.05), and the 100 most down-regulated transcripts yielded 8 candidate protective proteins (**Fig. 4A, 4G**).

**Figure 4.**
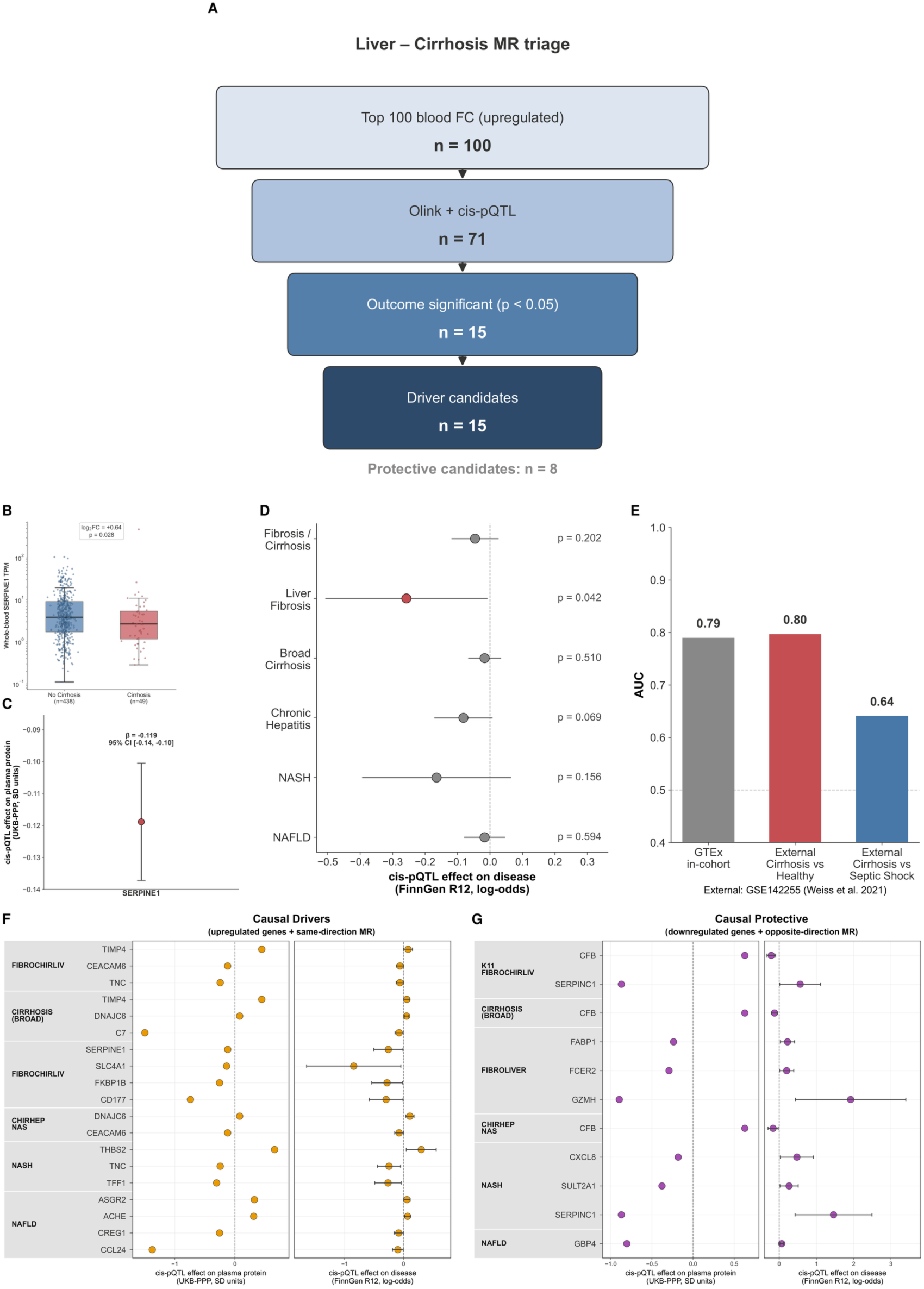
Mendelian randomization triages candidate causal plasma-protein drivers of liver-cirrhosis, illustrated for SERPINE1 and replicated in a living cohort. (A) MR triage funnel: the 100 most up-regulated blood transcripts, restricted to genes with both a UKB-PPP Olink measurement and a cis-pQTL instrument (n = 71), then filtered for nominal significance against a FinnGen liver phenotype (n = 15 drivers); the parallel opposite-direction pipeline yielded 8 protective candidates. (B) SERPINE1 whole-blood TPM in cirrhosis-positive (n = 49) versus cirrhosis-negative (n = 438) donors, with Mann-Whitney P. (C) SERPINE1 cis-pQTL effect on plasma protein (UKB-PPP, SD units), point and 95% CI. (D) SERPINE1 cis-pQTL effect across six FinnGen liver phenotypes, with the significant liver-fibrosis outcome highlighted. (E) External replication on GSE142255: in-cohort GTEx AUC, external cirrhosis-versus-healthy AUC, and cirrhosis-versus-septic-shock specificity control. (F, G) Candidate causal drivers (F) and protective proteins (G) ranked by minimum gene-phenotype MR P; left sub-axis, cis-pQTL effect on plasma protein; right sub-axis, effect on disease; rows grouped by FinnGen phenotype.

The top-ranked candidate drivers were: tenascin-C, thrombospondin-2, complement C7, the metalloproteinase inhibitor TIMP4, and the fibrinolysis regulator SERPINE1 (PAI-1) (**Fig. 4F**). These are matrix-remodeling and complement proteins with established causal roles in liver fibrosis. The triage therefore largely recovers known biology: it shows that the predictive blood signature is enriched for genuine causal drivers rather than incidental markers. SERPINE1 illustrates the full chain (**Fig. 4B-D**): its transcript is elevated in cirrhosis-positive donors (log2 fold change = +0.64, P = 0.028), its cis-pQTL lowers plasma SERPINE1 and lowers FinnGen liver-fibrosis risk (P = 0.042), so higher plasma SERPINE1 predicts higher risk, matching the profibrotic biology of PAI-1 [26].

Finally, we tested whether the signature holds outside the postmortem setting, where the circumstances of death cannot contribute. Applied to an independent whole-blood cohort of living patients (GSE142255 [27]; stable and decompensated cirrhosis, acute-on-chronic liver failure, healthy and septic-shock controls), the cirrhosis signature reached AUC = 0.80 against healthy controls, matching its in-cohort GTEx performance, and AUC = 0.64 against septic shock (**Fig. 4E**). The transfer to living patients is the strongest evidence that the signal reflects liver biology rather than agonal state, and the reduced but non-zero separation from septic shock indicates the signature is not merely the generic systemic-inflammation response shared by the two conditions.

The same triage applied to liver-steatosis, the second liver pathology to pass, nominated the collagen chaperone SERPINH1 (HSP47) and liver fatty-acid-binding protein FABP1 as anchor drivers, consistent with established roles in fibrosis [28] and NAFLD [29], with ADH1B and APOM among protective candidates (**Fig. S6A-C**; Supplementary Note 5).

## Methods

The Methods below implement the four steps of TRACE (**T**ranscriptomic **R**eading of **A**ge- associated organ pathologies and **C**ausal **E**valuation): donor-level pathology labeling (Methods 2), covariate-orthogonalized principal-component modeling and supervised prediction (Methods 3-4), pathway-level interpretation of the qualifying pairs (Methods 5), and Mendelian-randomization causal triage (Methods 6).

### 1. GTEx Whole-Blood Expression Data and Donor Metadata

Whole-blood gene expression, sample metadata, and donor pathology annotations were obtained from the GTEx v10 release Gene-level expression values were downloaded as transcripts per million (TPM) from the RNA-SeQC v2.4.2 pipeline and restricted to whole-blood samples. Donor age (in years), sex, race, Hardy scale, and total ischemic time were obtained from the restricted donor metadata file under approved dbGaP access. Tissue pathology annotations were obtained from the GTEx Histology Image Viewer, encoded by expert pathologists per slide. After matching donors with both whole-blood expression and at least one annotated postmortem tissue sample, the working dataset comprised 803 donors and 59,033 genes.

### 2. Donor-Level Pathology Labels and Cohort Definition

TRACE step 1, pathology labeling: For each tissue-pathology pair, donors were assigned a binary label from the matched tissue slide annotations. Raw pathology notes were parsed by a rule-based natural language pipeline that combined regular-expression matching against curated pathology vocabularies (iteratively expanded across versions by an LLM-assisted synonym-curation pass) with ConText-style negation handling [30]. Each candidate pathology assignment received a confidence score weighted by the presence of a positive qualifier and by multi-clause matches, and was zeroed when the matched phrase fell within the scope of a negation cue, except where the vocabulary entry was itself a negated term. Assignments below a pre-specified confidence threshold were discarded before label aggregation. A donor was labeled positive for a given pair if any slide from that tissue carried the target pathology term, and negative if at least one slide from the same tissue was annotated as showing no abnormalities; donors lacking either annotation were excluded. Pairs with fewer than five positive or five negative donors were also excluded. Filtering retained 53 pairs from the structured Pathology Categories field alone; NLP imputation of the free-text Pathology Notes field added six further pairs, yielding the final 59-pair pool across 20 tissue types. Pathology donor counts ranged from 108 to 633.

### 3. Cross-Validation Setup and Predictive Models

TRACE steps 2-3, covariate-orthogonalized modeling and prediction: The 20,000 most variable genes by per-gene TPM variance were retained for downstream modeling. A donor-level clinical covariates matrix was assembled from sex, age, race, Hardy scale, and total ischemic time, with median imputation applied within each training fold so that no test-fold information leaked into the imputation step. For each of the 59 tissue- pathology pairs, three nested random-forest classifiers [31] were fit under five-fold cross-validation, stratified by the binary label and grouped by donor identifier so that all samples from a donor remained in the same fold. The clinical covariates model used only the five donor variables. The gene expression model added the 100 genes ranked highest per training fold by absolute deviation from chance AUC. The PC + clinical covariates model replaced the gene-level features with the 100 principal components of highest absolute deviation from chance AUC, combined with the five donor variables. All random forests were trained with 500 trees, square-root feature subsampling at each split, balanced class weighting, and a fixed random seed (scikit-learn 1.9.0). Variance filtering, feature selection, PCA, and clinical-covariate median imputation were performed inside the training fold only, so that no information from held-out donors influenced model fitting. Performance was summarized by the pooled area under the receiver operating characteristic curve (AUC) across held-out folds. The added value of expression over clinical covariates was defined as ***AUC gain*** = ***AUC***(***PC*** + ***clinical covariates***) − ***AUC***(***clinical covariates***).

### 4. PC Model and Gene-Level Importance

To recover broader transcriptome structure while preserving interpretability at the gene level, a PC-based pipeline was run in parallel with the gene expression model. For each tissue-pathology pair and each cross-validation fold, the training-fold expression matrix was standardized and decomposed by principal-component analysis [32] into 800 components, which captured approximately 84.3% of total variance at the 100-component cap used downstream (**Fig. S1A)**. This choice follows the noise-reduction rationale established for high-dimensional aging biomarkers, where PC transformation integrates covariance across many features to separate systematic biological signal from feature-specific technical noise [33, 34]. Within each training fold, an orthogonality filter was applied to the 800 principal components before pathology-AUC selection. For each component, we computed its univariate AUC against each of the five clinical covariates (sex as a direct binary target; race and Hardy scale as one-versus-rest with the across-category maximum retained; age in years and total ischemic time binarized at the training-fold median) and dropped any component whose maximum AUC across the five covariates exceeded 0.70 (**Fig. S2A-B**). Of the surviving components, the 100 with the highest absolute deviation from chance AUC were combined with the five donor variables to train a random forest under the same hyperparameters as in Method 3. Tissue- pathology pairs meeting the joint criterion AUC ≥ 0.60 and AUC gain ≥ 0.05 over the clinical covariates model were retained for gene-level interpretation; four pairs met these criteria. To translate PC-level importance to genes, two normalization steps were applied per fold. First, random-forest Gini importances over the 100 selected components were normalized to sum to one across those components, yielding a per-component importance weight **w(p)**. Second, the absolute PCA loadings of each selected component were normalized across genes, yielding a per-gene share **ℓ(p, g)** of that component’s expression footprint. Gene-level importance was then computed by averaging the weighted shares across folds:

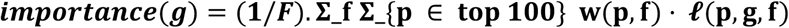

where F = 5 is the number of folds and the inner sum runs over the 100 components selected for that fold. Entries that remained unannotated transcript identifiers (ENSG- prefixed, primarily pseudogenes and predicted non-coding loci) were dropped at this step so that downstream pathway and Mendelian randomization analyses operated on a symbol-keyed gene universe of approximately 16,674 genes per pair (representing ∼83% of the 20,000-gene variance-filtered set). Cross-model concordance with the direct gene- RF ranking was quantified by Spearman’s rank correlation per pair.

### 5. Pathway Enrichment Analysis

Because the PC-derived gene importance scores spread signal across the full transcriptome, pathway enrichment was performed on the complete Pathology gene ranking using pre-ranked gene set enrichment analysis, implemented in GSEApy v1.2.1 [35] with 1,000 permutations and gene-set size constraints from 10 to 500, seeded from the same random seed as the predictive models. Two pathway libraries were queried: KEGG 2026 gene-set library and the Reactome Pathways 2024 gene-set library, both retrieved from Enrichr via GSEApy. Pathways with FDR q-value < 0.05 and normalized enrichment score (NES) > 0 were retained for interpretation; supplementary heatmaps show pathways at FDR < 0.25 for broader visual context (**Fig. S5C**). The unsigned nature of PC-derived gene importance means only positive-NES pathways carry biological interpretation: pathways enriched at the bottom of the ranking correspond to the uninformative null tail and are not reported. For each qualifying tissue-pathology pair and each library, the top-ranked pathways by FDR were retained for biological interpretation.

### 6. Two-Sample Mendelian Randomization

TRACE step 4, causal triage: Causal candidate identification for the qualifying Liver tissue- pathology pairs used two-sample Wald-ratio Mendelian randomization. Plasma protein exposure instruments were the lead cis-pQTLs listed in Supplementary Table 9 of Sun et al. 2023, which reports 14,287 primary pQTLs (12,332 trans, 1,955 cis) reaching the study-wide discovery threshold of P < 1.7 × 10⁻¹¹ in the UKB-PPP discovery cohort. We restricted to the 1,955 cis-pQTLs, spanning 1,954 unique proteins, and retained one lead cis-pQTL per protein. For each top-ranked blood transcript with a UKB-PPP Olink measurement, we extracted the cis-pQTL rsID and its discovery effect estimate and standard error with respect to the effect allele. Disease outcome summary statistics were obtained from FinnGen R12 for six liver phenotypes (NAFLD, NASH, CIRRHOSIS BROAD, K11 FIBROCHIRLIV, CHIRHEP NAS, FIBROLIV). FinnGen alleles were harmonized to the UKB-PPP effect allele by simple two-allele match: if the FinnGen alternate allele equalled the UKB-PPP effect allele, the FinnGen beta was kept as-is; otherwise it was negated. Palindromic variants were not further reconciled using allele-frequency information.

The Wald-ratio MR was computed using standard mendelian randomization process. Two complementary direction filters were applied. A same-direction filter, in which the cis-pQTL effect on plasma protein and the cis-pQTL effect on disease have matching signs, was applied to the 100 most upregulated transcripts to identify candidate causal drivers. An opposite-direction filter was applied to the 100 most downregulated transcripts to identify candidate causal protective proteins. Gene-phenotype MR results were retained at nominal MR p-value < 0.05 for candidate prioritization; no genome-wide multiple-testing correction was applied at this stage. For each gene, the minimum p across the six FinnGen R12 liver phenotypes was used to rank candidates; no correction for the pooled testing across these phenotypes was applied. All cis-pQTL lookups and FinnGen R12 variant queries were cached as per-phenotype Parquet files for reproducibility.

### 7. External-Cohort Validation of the Liver-Cirrhosis Signature

To test whether the liver-cirrhosis signature generalizes beyond the postmortem GTEx cohort, we applied it to an independent whole-blood transcriptome dataset of living patients (GEO accession GSE142255), comprising 46 samples: 31 with cirrhosis of any stage and 15 without (7 healthy controls and 8 patients with septic shock but no cirrhosis). Expression was profiled on the Affymetrix Human Transcriptome Array 2.0 (platform GPL17586); probe-level log2 intensities were mapped to gene symbols using the GPL17586 annotation, aggregating multiple probes per gene by their mean to yield a gene-by-sample matrix.

The signature was defined as the GTEx liver-cirrhosis gene ranking obtained by back-projecting principal-component importances to genes (Methods 4), taking the top-K genes by importance for K = 50, 100, and 200. For each K, the signature genes present in the external matrix (35, 63, and 107 genes, respectively) were retained; each gene’s expression was standardized across the 46 samples, and a per-sample signature score was computed as the importance-weighted sum of these standardized values. Because principal-component-derived importances are unsigned, the scores carry no intrinsic direction; classification performance was therefore summarized by a direction-agnostic area under the ROC curve, AUC = max(AUC, 1 − AUC), and per-sample scores were sign-aligned to the case label for visualization. Discrimination was evaluated three ways: against all 15 non-cirrhotic samples, against healthy controls only, and against septic-shock samples only (a specificity control for generic systemic inflammation). We report the K = 100 result in the main text; performance was stable across K (**Fig. 4E**).

## Discussion

Across 59 GTEx tissue-pathology pairs, the whole-blood transcriptome carried confounder-independent signal for four, and for one, liver-cirrhosis, the signal was strong (AUC = 0.79; gain = +0.215 over a covariate baseline), replicated in living patients, specific against systemic inflammation, and enriched for causal fibrosis drivers on genetic triage. The broader reading is deliberately modest: for most organ pathologies in this benchmark, the whole-blood transcriptome does not carry a trace that survives donor demographics and the circumstances of death, and cirrhosis is the clear exception rather than the rule.

The framework makes two claims. The first is methodological: a blood signal earns interpretation only after it clears donor demographics and procedural variance, which we enforce by benchmarking every model against a covariate baseline and orthogonalizing the component pool against all five covariates per fold (Methods 4). The second is causal: the surviving signal can be triaged toward the features more likely to drive the pathology, separating this work from association-only blood-biomarker studies. That reach should not be overstated, as our strongest candidates are proteins already known to drive fibrosis, so the triage is best read as confirming that the predictive signature is causally enriched rather than as discovering new causal biomarkers. The deepest caveats to both claims, residual death-mode confounding and single-instrument Mendelian randomization, are detailed below.

### Limitations

First, GTEx is a postmortem cohort in which the circumstances of death dominate blood-expression variance. Our controls reduce this confound but cannot remove it, and generalization of GTEx-trained classifiers to living patients is tested here only for the strongest pair; comparable living-cohort data for the other three qualifying pairs are limited or unavailable. Second, the cirrhosis signature (AUC = 0.79) does not match established clinical tests for liver fibrosis, which reach higher discrimination from routine blood chemistry. TRACE is therefore a framework and a resource, not a diagnostic; its value is in defining where blood carries organ-pathology signal and in triaging that signal toward mechanism, not in outperforming existing assays.

Third, the PC-derived gene rankings are correlative and unsigned: a high rank reflects loading magnitude on informative components, not a measured effect on pathology, and a fraction of the top transcripts are non-coding. The rankings are hypothesis-generating, and pathway enrichments identify features concentrated among informative components rather than genes up- or down-regulated in disease. Fourth, the Mendelian-randomization triage uses single-instrument Wald-ratio tests at nominal significance, without colocalization or multiple-testing correction. It cannot distinguish a shared causal variant from linkage, or vertical from horizontal pleiotropy. The nominated candidates should be treated as a prioritized list awaiting colocalization, multiple-instrument inverse-variance-weighted estimation with pleiotropy-robust sensitivity analyses [36, 37], and replication across pQTL platforms.

By making explicit which organ pathologies do and do not leave a confounder-independent trace in blood, and by triaging that trace toward causal biology, TRACE offers a template for the coming wave of biobank-scale studies in which blood is the only accessible tissue, and a reminder that, for most organ pathologies, reading the organ from blood remains harder than it looks.

## Code availability

All development code is available at https://github.com/Cranjit9/TRACE. The pipeline (notebooks NB01–NB17 and the trace_path package) reproduces the tables and figures reported here from the input data listed above.

## Data availability

All datasets analysed in this study are publicly available. GTEx v10 whole-blood gene expression, sample attributes, and histopathology annotations were obtained from the GTEx Portal (https://gtexportal.org); restricted donor metadata (age and total ischemic time) were accessed under an approved dbGaP application (accession phs000424). Plasma cis-pQTL instruments were obtained from the UK Biobank Pharma Proteomics Project (Supplementary Table 9 of Sun et al.). Liver-disease outcome summary statistics were obtained from FinnGen Release 12 (https://www.finngen.fi). The external validation cohort is available from the Gene Expression Omnibus under accession GSE142255, with array annotation from platform GPL17586. The UK Biobank Pharma Proteomics Project, FinnGen, and the GSE142255 cohort were used as publicly available, de-identified summary or aggregate data from studies with their own ethical approvals and participant consent.

## Acknowledgement

This work was supported by Dr. Sanju Sinha’s start-up funding from Sanford Burnham Prebys Medical Discovery Institute.

## Author contributions

S.S. conceived and designed the TRACE framework. R.K.S. developed the computational framework, performed the data analysis and model validation, and contributed to writing the manuscript. K.A. contributed to figure preparation, manuscript review, and independent reproduction of the analysis pipeline. S.S. supervised the project and mentored manuscript writing. All authors read and approved the final version of the manuscript.

