## Supplementary material for "Whole-blood transcriptomic traces of organ pathology and their causal triage": Supp_Figures_and_Text

### Supplementary Notes and Figures

#### Supplementary Notes

1. Natural-language rescue of free-text notes expanded the screen from 53 to 59 pairs. GTEx's structured Pathology Categories field alone yielded 53 tissue-pathology pairs meeting the minimum class-size threshold. A regular-expression pipeline with ConText-style negation handling recovered six further pairs from the free-text Pathology Notes field, bringing the screened pool to 59(**Fig. 2A**). The qualifying rate (4 of 59) is unchanged whether or not the imputed pairs are included, so the rescue adds coverage breadth rather than qualifying calls.

2. Cross-validation stability supports the qualifying calls. Per-fold AUC standard deviation across the 59-pair pool ranged from 0.02 to 0.16, and all four qualifying pairs fell below the median fold variance (**Fig. S2E**). Liver-cirrhosis was the most stable fold-to-fold despite its small positive class (n = 49). For pairs at the low end of the sample-size range, fold variance is non-trivial, so we report mean and standard deviation across folds for every pair.

3. The clinical-covariate baseline isolates blood-expression-specific signal. Total ischemic time dominates the principal-component structure of the whole-blood matrix (**Fig. S1B**). For the four qualifying pairs, the expression-plus-covariates model exceeds both the covariates-only and the expression-only model (**Fig. S3A**), confirming that the gain attributed to blood expression is reproducible whether or not the model also has access to the covariates.

4. Principal-component back-projection agrees with gene-level random-forest importance. The PC-derived gene ranking was concordant with an independent gene-level random forest on the same donors at Spearman rho = 0.56 (liver-cirrhosis), 0.55 (small intestine-nodularity), 0.25 (liver-steatosis), and 0.23 (lung-congestion) (**Fig. S4A-C**). Agreement was highest in the two largest cohorts, consistent with back-projection recovering the features the gene-level model weights when sample size permits.

5. The causal triage for liver-steatosis parallels liver-cirrhosis. The same two-sample Mendelian-randomization pipeline was applied to liver-steatosis, the second liver pathology to clear the gate. Same-direction MR on the top-100 up-regulated transcripts yielded 60 cis-pQTL-anchored gene tests, 7 nominally significant ($P < 0.05$) against at least one FinnGen phenotype (**Fig. S6B**); opposite-direction MR on the top-100 down-regulated transcripts yielded 75 tests with 12 nominally significant protective candidates (**Fig. S6C**). The driver panel is anchored by SERPINH1 (HSP47) and FABP1, consistent with roles in hepatic fibrosis and lipid handling, and the protective panel by ADH1B and APOM, consistent with roles in alcohol metabolism and lipid transport. As in the main analysis, these are nominal single-instrument candidates for follow-up, not established causal drivers.

#### Supplementary Figures

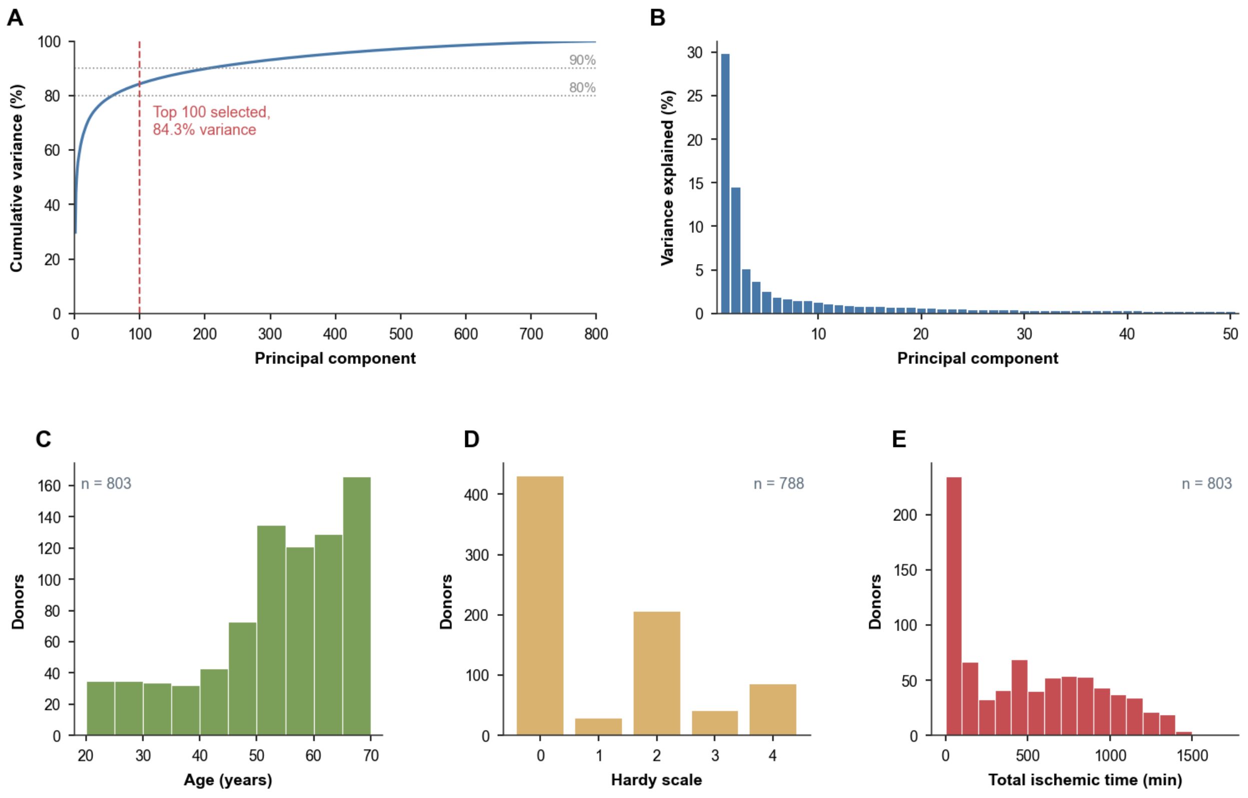


***Figure S1***. *GTEx v10 whole-blood cohort structure and PCA variance decomposition. (A) Cumulative variance across all 800 principal components of the top-20,000 variance-filtered whole-blood transcriptome; the dashed line marks the top-100 PCs carried forward. (B) Per-PC variance explained (scree) for the top-50 PCs. (C) Age distribution of the 803 donors. (D) Hardy-scale distribution. (E) Total-ischemic-time distribution.*

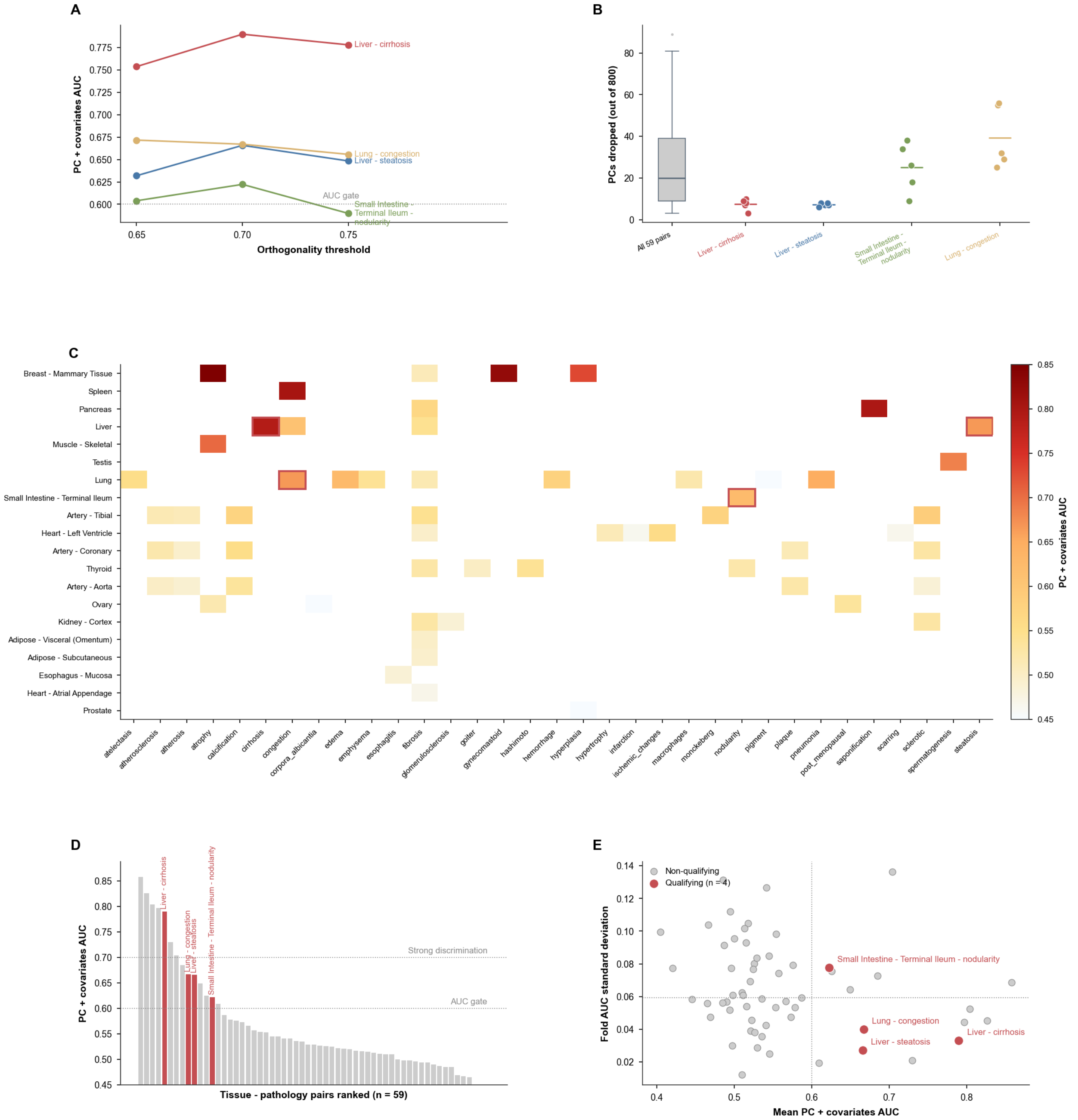


***Figure S2***. *Screening pipeline: covariate-orthogonal PC filter and the full 59-pair landscape. (A) PC-plus-covariates AUC of the four qualifying pairs across three orthogonality thresholds (0.65, 0.70, 0.75); the dashed line marks the AUC gate at 0.60. (B) Distribution of PCs dropped per fold at the 0.70 threshold, for all 59 pairs and for each qualifying pair. (C) Tissue-by-pathology AUC heatmap across all 59 pairs; coral outlines mark the four qualifying pairs. (D) Ranked PC-plus-covariates AUC across the 59 pairs, qualifying pairs highlighted, against the AUC gate (0.60) and a strong-discrimination reference (0.70). (E) Fold stability: mean AUC versus cross-fold standard deviation, qualifying pairs annotated.*

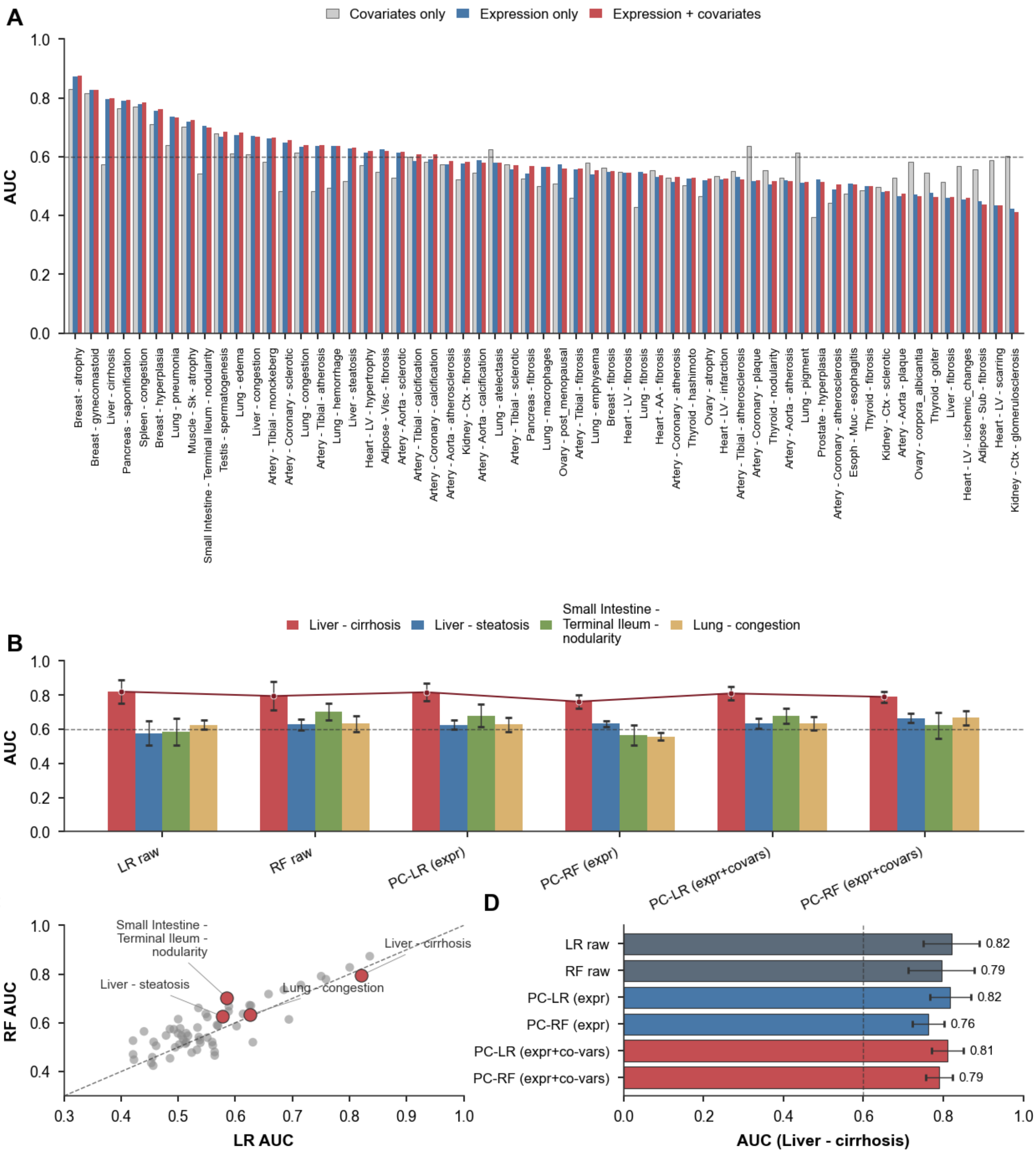


***Figure S3***. *Model comparison across covariates-only, expression-only, and combined predictors. (A) Three-way AUC across all 59 pairs: covariates-only, expression-only, and expression-plus-covariates; the dashed line marks the AUC gate at 0.60. (B) Six-model AUC for the four qualifying pairs, with liver-cirrhosis traced across models. (C) Logistic-regression versus random-forest AUC across the 59 pairs, qualifying pairs annotated. (D) Six-model AUC for liver-cirrhosis.*

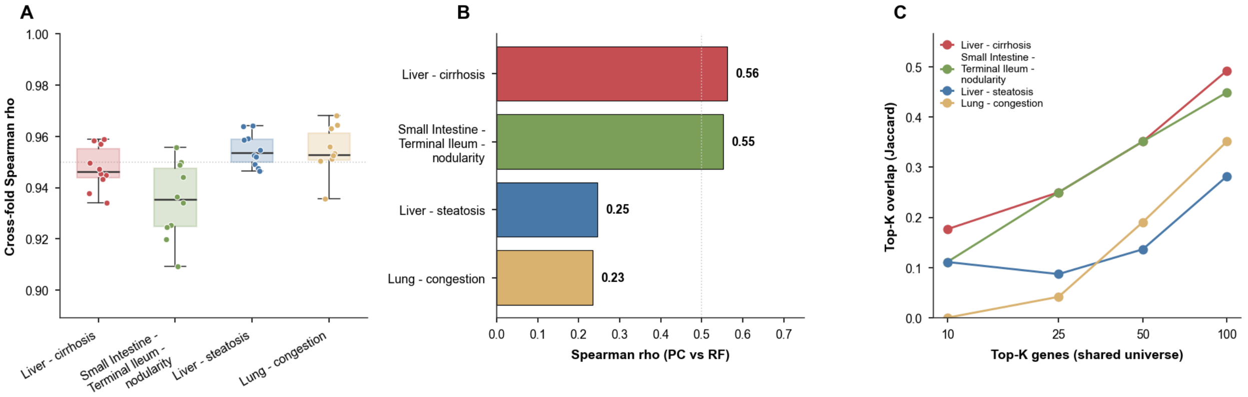


***Figure S4.*** *Principal-component back-projection is stable across folds and concordant with gene-level importance. (A) Cross-fold Spearman rho of PC-derived gene rankings across the four qualifying pairs. (B) Spearman rho between PC-derived and gene-level random-forest importance for each qualifying pair. (C) Top-K overlap (Jaccard) between the two rankings at K = 10, 25, 50, and 100, within the shared gene universe.*

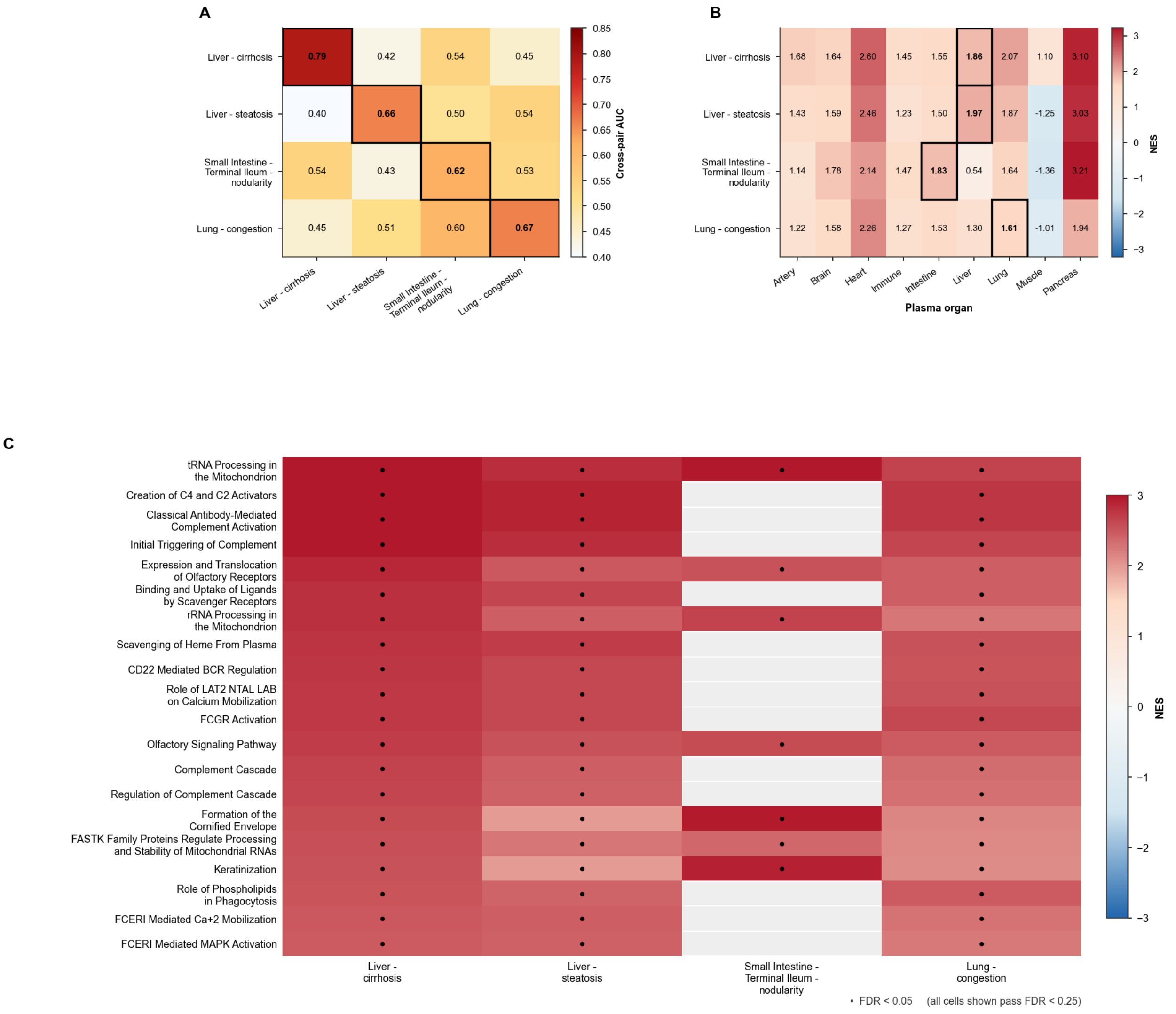

***Figure S5.*** *Biological interpretation of the four qualifying signatures. (A) Signature-specificity matrix: AUC of each pair's PC signature applied to every other pair; diagonal (own-pair) cells outlined. (B) Cross-organ plasma enrichment (NES) using the plasma-organ atlas of Oh et al. [3]; own-organ cells outlined. (C) Reactome 2024 top-20 pathway union across the four qualifying pairs, ordered by liver-cirrhosis NES; dots mark FDR < 0.05, and all shown cells pass FDR < 0.25.*

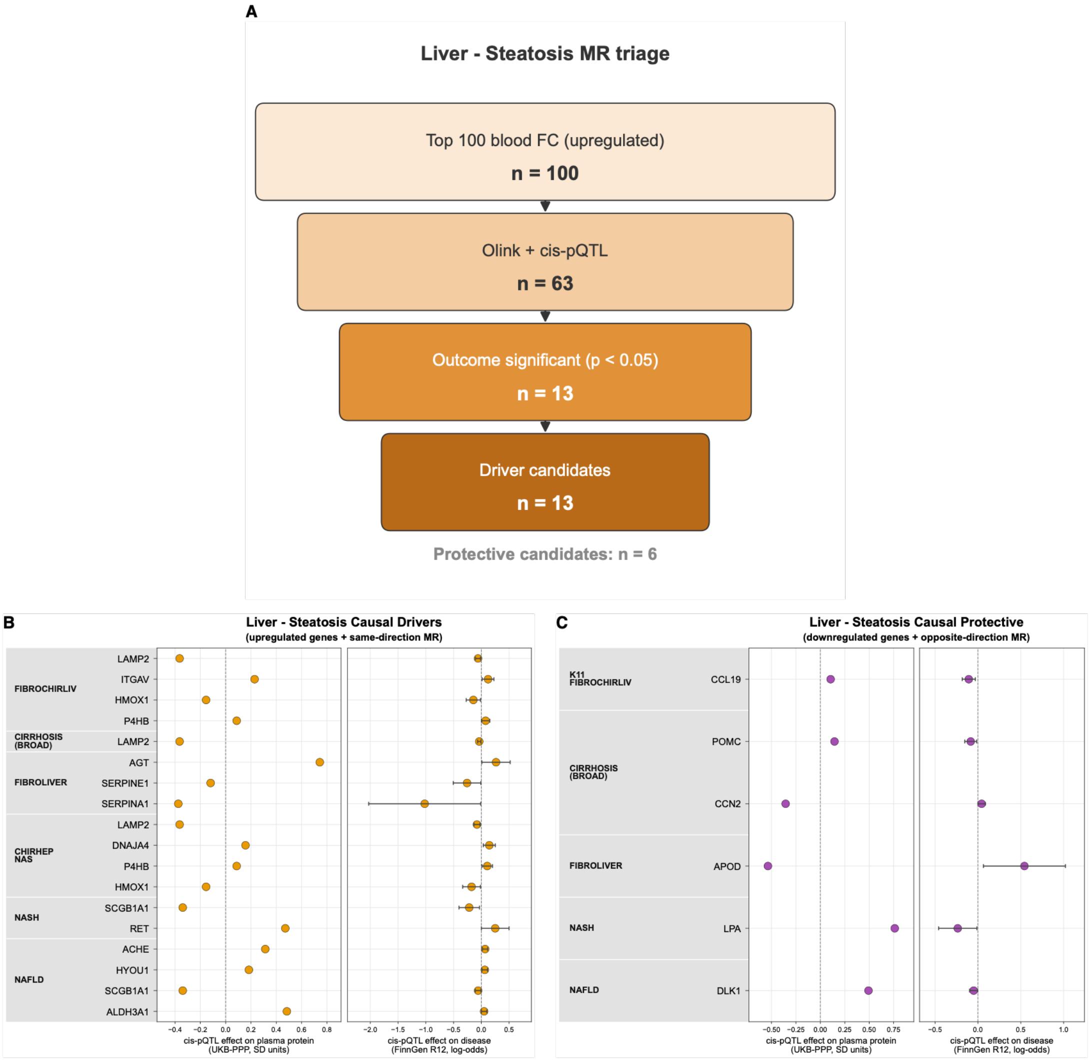


***Figure S6.*** *Causal triage for liver-steatosis. (A) MR funnel: the top-100 up-regulated transcripts filtered through Olink and cis-pQTL coverage and outcome significance (P < 0.05) to concordant-sign driver candidates, with the protective count reported below. (B) Driver-candidate forest: cis-pQTL effect on disease log-odds (UKB-PPP and FinnGen R12); error bars are 95% confidence intervals. (C) Protective-candidate forest for transcripts down-regulated in blood whose plasma-protein direction is protective against the FinnGen phenotype.*
